# Implications of cranial arterial stenosis and dolichoectasia for cerebral small vessel disease etiopathogenesis: findings from a prospective mild stroke cohort

**DOI:** 10.64898/2026.01.26.26344896

**Authors:** Fei Han, Una Clancy, Carmen Arteaga-Reyes, Michael J. Thrippleton, Maria Del C. Valdés Hernández, Daniela Jaime Garcia, Michael S. Stringer, Ellen Backhouse, Francesca M. Chappell, Yajun Cheng, Dillys Xiaodi Liu, Junfang Zhang, Angela C.C. Jochems, Eleni Sakka, Charlotte Jardine, Gayle Barclay, Donna McIntyre, Iona Hamilton, Rosalind Brown, Yi-Cheng Zhu, Fergus N. Doubal, Joanna M. Wardlaw

## Abstract

**Background:** Stenosis and dolichoectasia of cranial arteries likely reflect distinct mechanisms. Their contributions to lacunar stroke and cerebral small vessel disease (cSVD) remain contentious. We investigated associations of large artery stenosis (LAS) and arterial widening with stroke subtype, cSVD markers, incident infarcts, and clinical outcomes.

**Methods:** We prospectively recruited patients with lacunar or mild non-lacunar stroke, with demographic, stroke-related, cognitive, functional, and MRI (index and incident infarcts, cSVD markers) assessments at baseline and one year. LAS was defined as ≥50% intracranial or cervical artery stenosis; basilar artery dolichoectasia (BADE) by basilar artery diameter, bifurcation height, and lateral displacement; and intracranial carotid and middle cerebral artery diameters were also measured. Associations were estimated using multivariable regression adjusted for age, sex, and vascular risk factors. We further conducted a systematic literature review to synthesize evidence on relationships between large artery pathology and cSVD.

**Results:** Among 229 patients (mean age 65.9±11.1 years; 131 [57.2%] lacunar stroke), LAS and BADE were present in 20.5% and 15.7%, respectively. After adjustment, LAS (odds ratio [OR], 0.49; 95%CI, 0.23-0.99) and the presence of any embolic source were associated with lower odds of lacunar versus non-lacunar stroke, and not with cSVD markers or incident infarcts. In contrast, BADE was strongly associated with lacunar stroke (OR, 4.67; 95%CI, 1.87-13.14), higher cSVD scores (ordinal analysis; OR, 2.57; 95%CI, 1.28-5.25), incident infarcts (75% subcortical; OR, 2.29; 95%CI, 1.01-5.14), and greater progression of white matter hyperintensities over one year (β, 0.15; 95%CI, 0.01-0.29; per log_10_-transformed volume). Similar associations were observed for wider intracranial arteries. The systematic review supported these findings.

**Conclusions:** cSVD, including lacunar stroke, was unrelated to LAS, but strongly associated with dolichoectasia and wider arteries. These findings support a non-atheromatous, intrinsic microvascular pathology, particularly segmental arteriolar disorganization, as the principal mechanism of lacunar stroke and cSVD. Mechanism-specific diagnostic and therapeutic strategies are warranted.

**Clinical Perspective:** *What Is New?:*

● Large artery stenosis was unlikely to represent a causal mechanism for lacunar stroke and showed no association with cerebral small vessel disease (cSVD) imaging markers.
● Dolichoectasia and intracranial arterial widening emerged as vascular phenotypes strongly associated with cSVD, including its progression and lacunar stroke subtype.

*What Are the Clinical Implications?:*

● Distinct large artery phenotypes have divergent etiopathological implications for cSVD. Our findings support a non-atheromatous, intrinsic microvascular pathology as the principal mechanism of lacunar stroke and cSVD.
● Mechanism-based therapeutic strategies for lacunar stroke and cSVD, moving beyond conventional approaches focused on atherosclerosis or cardioembolism, are warranted.

## Introduction

Cranial artery stenosis and dolichoectasia likely represent distinct forms of large artery pathology. Large artery stenosis (LAS) arises from focal luminal narrowing due to atherosclerotic plaque,^1^ whereas dolichoectasia is generally considered to reflect non-atherosclerotic dilatation and tortuosity.^2^ These conditions differ in morphology and are thought to involve distinct pathophysiological processes, with potentially different impacts on and clinical implications for downstream small vessels and stroke subtypes. Although associations between large artery disease and cerebral small vessel disease (cSVD) have been reported,^3–5^ most studies were cross-sectional and rarely investigated both stenosis and dolichoectasia within well-phenotyped stroke cohorts with longitudinal follow-up.

Lacunar strokes, accounting for 20-30% of ischemic strokes,^6^ are mainly attributed to intrinsic cSVD, particularly the ‘segmental arterial disorganization’ described in Fisher’s seminal clinicopathological studies.^7^ In hypertensive patients with lacunar stroke, pathological studies have documented arteriolar expansion, mural cellular and fibrinoid infiltration with wall destruction, and luminal occlusion by fibrinoid material. However, the relative contributions of atherothromboembolism, cardioembolism, and intrinsic microarteriopathy remain debated. Approximately 15% of lacunar stroke patients harbor potential embolic sources, including large artery atheroma, but their causal relevance is uncertain.^8–11^

In this prospective study of lacunar stroke, with non-lacunar ischemic strokes of similar severity as comparators, we investigated how atheromatous stenosis, dolichoectasia and arterial widening relate to stroke subtypes, cSVD and its progression, longitudinal incident infarcts and clinical outcomes. By directly comparing these two large artery phenotypes within the same population, we aimed to clarify their respective roles in cSVD and the pathogenesis of lacunar stroke, with potential implications for targeted management.

## Methods

### Data Availability

The data supporting the findings of this study are available within the article and Supplemental Material. All other deidentified participant data can be made available on reasonable request to the corresponding author.

## Study design and participants

The Mild Stroke Study 3 is a prospective cohort of patients with recent lacunar or mild non-lacunar ischemic stroke, consecutively recruited from Edinburgh stroke services between 2018 and 2021. The detailed protocol has been published.^12^ Mild stroke was defined as National Institutes of Health Stroke Scale <8 at initial stroke presentation and modified Rankin scale (mRS) ≤2 at enrollment (between 1 and 3 months post-stroke). Exclusion criteria included contraindications to MRI and severe neurological, cardiac, or respiratory comorbidities. All underwent standard guideline-based stroke investigations and secondary prevention. Clinical and MRI assessments were conducted at baseline and at 6 and 12 months, with additional MRI scans at 3 or 9 months for those with lacunar stroke or moderate-to-severe white matter hyperintensities (WMH) (Figure 1).

**Figure 1.**
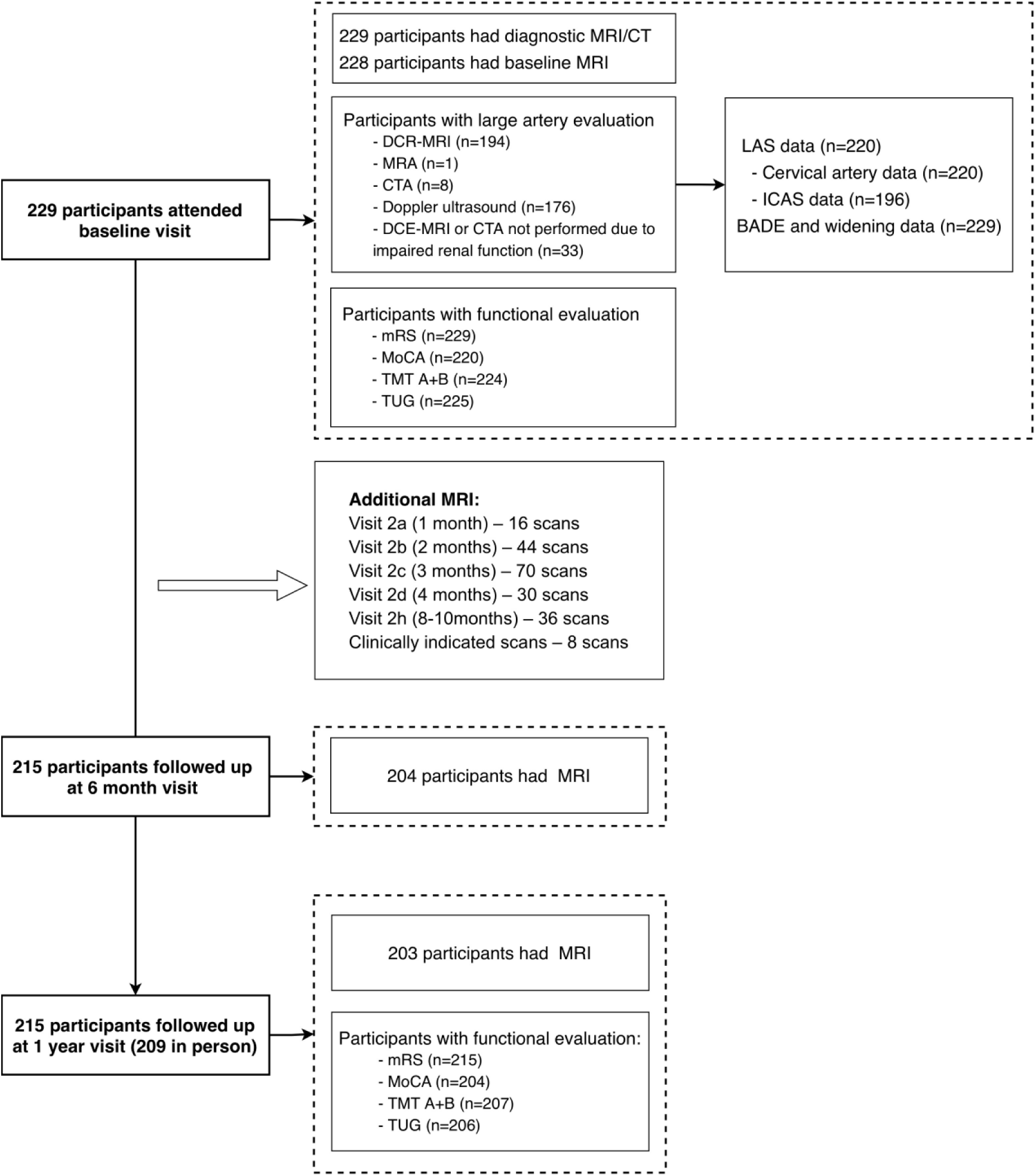
Flowchart of the study. Abbreviations: LAS, large artery stenosis; BADE, basilar artery dolichoectasia; DCE-MRI, dynamic contrast-enhanced magnetic resonance imaging; MRA, magnetic resonance angiography; CTA: computed tomography angiography; mRS, modified Rankin score; MoCA, Montreal Cognitive Assessment; TMT, Trail Making Test; TUG, Timed Up-and-Go.

The study was approved by the Southeast Scotland Regional Ethics Committee (Ref.18/SS/0044), conducted under the Declaration of Helsinki, and registered under ISRCTN 12113543. All participants provided written informed consent.

## Clinical measurements

The presenting (‘index’) strokes were subtyped clinically as lacunar or non-lacunar stroke syndrome according to the Oxfordshire Community Stroke Project classification,^13^ with a corresponding recent infarct on diagnostic MRI or CT, or if no visible infarct, no other explanation for the stroke symptoms. When clinical and imaging classifications differed, the imaging lesion defined the final subtype.^14^ Demographics, vascular risk factors, and cardiovascular comorbidities were recorded. Premorbid intelligence was assessed with the National Adult Reading Test (NART).^15^ Functional status (mRS), cognition (Montreal Cognitive Assessment [MoCA]^16^, Trail Making Test [TMT] B/A ratio^17^), and mobility (Timed Up-and-Go [TUG]) were evaluated at baseline and one year. Recurrent stroke or transient ischemic attack (TIA) was diagnosed using established criteria and confirmed by stroke specialists.

## Imaging Acquisition

All participants underwent diagnostic MRI or CT at stroke presentation. Baseline and follow-up cerebrovascular imaging were performed using one dedicated 3T research scanner (Siemens Prisma; Siemens Healthcare, Erlangen, Germany). Sequences included 3D T1-weighted, T2-weighted, fluid attenuated inversion recovery (FLAIR), susceptibility-weighted, and diffusion-weighted imaging (DWI), with additional advanced protocols. At baseline, blood-brain barrier integrity was assessed using dynamic contrast-enhanced (DCE)-MRI with intravenous gadobutrol (0·1mmol/kg, Gadovist 1M; Bayer, Germany), unless estimated glomerular filtration rate was <30 ml/min. Detailed MRI parameters are provided in Supplemental Methods and published protocol.^12,18^

## Imaging Analysis

All imaging analyses were performed blinded to clinical data and supervised by a neuroradiologist (J.M.W.). Structural MRI and large artery assessments were conducted independently by different raters at separate time points, each blinded to the other.

### Index and incident infarcts

‘Index’ and ‘incident’ infarcts were defined as previously described.^18^ The index infarct was confirmed as an acute lesion on diagnostic imaging consistent with the presenting stroke syndrome. Radiologically detected incident infarcts were new infarcts between index stroke and 1-year follow-up, visible on DWI, FLAIR, and/or T2-weighted sequences, and absent on prior scans. All infarcts were visually assessed on each diagnostic and study scan using a standardized proforma.^18^ Lesions were classified as subcortical or cortical; subcortical infarcts were further categorized by shape as round/ovoid or tubular.^19^

### Cerebral small vessel disease markers

cSVD markers were assessed according to STRIVE-2 criteria.^19^ Lacunes and microbleeds were counted, and a summary cSVD score was constructed.^20^ Cerebral atrophy was rated using validated scales.^21^ Quantitative MRI analyses applied semi-automated pipelines to segment and quantify intracranial volume (ICV), normal-appearing white matter, WMHs, perivascular spaces (PVS), and infarct volumes.^22,23^ WMH progression was defined as baseline normal-appearing white matter converting to WMH at one year. Brain volume, WMH and infarct volumes were normalized to ICV (%ICV), whereas basal ganglia and centrum semiovale PVS to the corresponding volumes of region of interest (%ROIV).

### Large artery stenosis and dolichoectasia

As MR angiography was not routinely performed, intracranial atherosclerotic stenosis (ICAS) was assessed on arterial-phase DCE-MRI, reviewed in multiple planes and graded at the narrowest lumen using established criteria (Figure S1).^24^ Intracranial arteries assessed included internal carotid arteries (ICA), middle cerebral arteries (MCA), anterior and posterior cerebral arteries, vertebral arteries, and basilar artery (BA). When stenosis was suspected, structural sequences were reviewed for confirmation. Cervical ICA stenosis was further assessed by ultrasound and graded by the North American Symptomatic Carotid Endarterectomy Trial criteria.^25^ Where available, MR or CT angiography was also reviewed. ICAS was defined as ≥50% stenosis in any intracranial artery; LAS was defined as ICAS or ≥50% cervical artery stenosis. A potential embolic source included LAS or major cardiac abnormalities (atrial fibrillation, patent foramen ovale, ischemic heart disease, valvular disease, or heart failure).

Maximum diameters of the BA, bilateral ICAs (vertical cavernous segment), and MCAs (M1 segment) were measured on axial T2-weighted images to 0.01mm (Figure S2). In stenotic cases, the least affected segment was measured. Diameters were adjusted for ICV using residual correction, and bilateral ICA/MCA diameters were averaged. BA bifurcation height and lateral displacement were rated using a validated four-point scale (Figure S2).^26^ Bifurcation height was scored as: 0=at/below dorsum sellae, 1=within suprasellar cistern, 2=at the level of third ventricle floor, 3=indenting and elevating third ventricle floor. Lateral displacement was graded by maximal lateral deviation: 0=midline, 1=medial to clivus or dorsum sellae margin, 2=lateral to this margin, 3=cerebellopontine angle cistern. Basilar artery dolichoectasia (BADE) was defined as BA diameter >4.5mm, bifurcation height ≥2, or lateral displacement ≥2.^26^

Intra-rater reliability showed intraclass correlation coefficients for diameters were 0.95 (BA), 0.85 (ICA), and 0.9 (MCA). Weighted kappa was 0.81 for ICAS, and 0.85 for BADE.

## Systematic review

We systematically reviewed studies investigating associations between large artery pathology and lacunar stroke or cSVD. MEDLINE was searched through Jun 22, 2025. Random-effects meta-analyses were performed to estimate the prevalence and laterality of LAS in lacunar stroke. The full search strategy, selection criteria, and methodological details are provided in the Supplemental Methods.

## Statistical analysis

Data are presented as mean (standard deviation), median (interquartile range), or count (%). Group comparisons used χ² tests for categorical and *t* or Mann-Whitney *U* tests for continuous variables. Infarct analyses were conducted at both per-person and per-infarct levels. Per-person analyses applied multivariable regression adjusting for age, sex, smoking, hypertension, diabetes mellitus, hyperlipidemia, and body mass index to assess associations of LAS and BADE with index stroke subtype, number and volume of index infarcts, incident infarcts, and recurrent stroke or TIA. Per-infarct analyses used Rao-Scott cluster-adjusted χ² tests to account for multiple infarcts per patient, comparing infarct subtype with LAS, BADE, or embolic sources. Among patients with LAS, infarct location relative to the stenotic artery was also examined.

Associations of LAS and BADE with cSVD markers at baseline and one year were analyzed using multivariable linear or ordinal logistic regression, with the proportional odds assumption verified by the Brant test. Models were adjusted for age, sex, and vascular risk factors. WMH and PVS volumes were log_10_-transformed. Lacune count was included in models of atrophy, and one-year outcomes were additionally adjusted for baseline cSVD score. Results are reported as odds ratios (OR) or regression coefficients with 95% CIs. Clinical outcomes (mRS, MoCA, TMT B/A ratio, and TUG) were analyzed with logistic or linear regression, adjusting for age, sex, baseline cSVD score, and NART for cognitive measures.

Sensitivity analyses tested the robustness of the main findings in index and incident infarct models by (1) examining ICAS, any potential embolic source, and atrial fibrillation instead of LAS, and (2) adjusting for baseline cSVD score. For analyses focusing on cSVD markers, sensitivity analyses further included (3) examining ICAS, any potential embolic source, and atrial fibrillation instead of LAS, and substituting BADE with continuous arterial diameters (ICA, MCA, and BA). In models where arterial diameters were analyzed as continuous exposures, LAS status was included as an additional covariate. (4) Multiplicative interaction terms between LAS and BADE were included in the above models to assess potential effect modification. All analyses were performed using R (version 4.4.2), with two-sided *P*<0.05 considered significant.

## Results

### Baseline characteristics

We recruited 229 participants (mean age, 65.9±11.1 years; 152 [66.4%] male; Table 1), including 131 (57.2%) with lacunar and 98 (42.8%) with non-lacunar stroke. Most participants (n=168) had a single index infarct, 25 had two and 20 had three or more. LAS was identified in 47 (20.5%) participants, comprising 40 (17.4%) with ICAS, 13 with cervical ICA stenosis, and 6 with both. A potential embolic source was present in 97 (42.4%) participants, of whom 23 had atrial fibrillation, and BADE in 36 (15.7%). Details of stenosis location and arterial diameters are presented in Table S1 and S2. At baseline, 204 (89.1%) participants received antiplatelet therapy, 26 (11.4%) received anticoagulation, and 211 (92.1%) were treated with lipid-lowering medications.

**Table 1.**
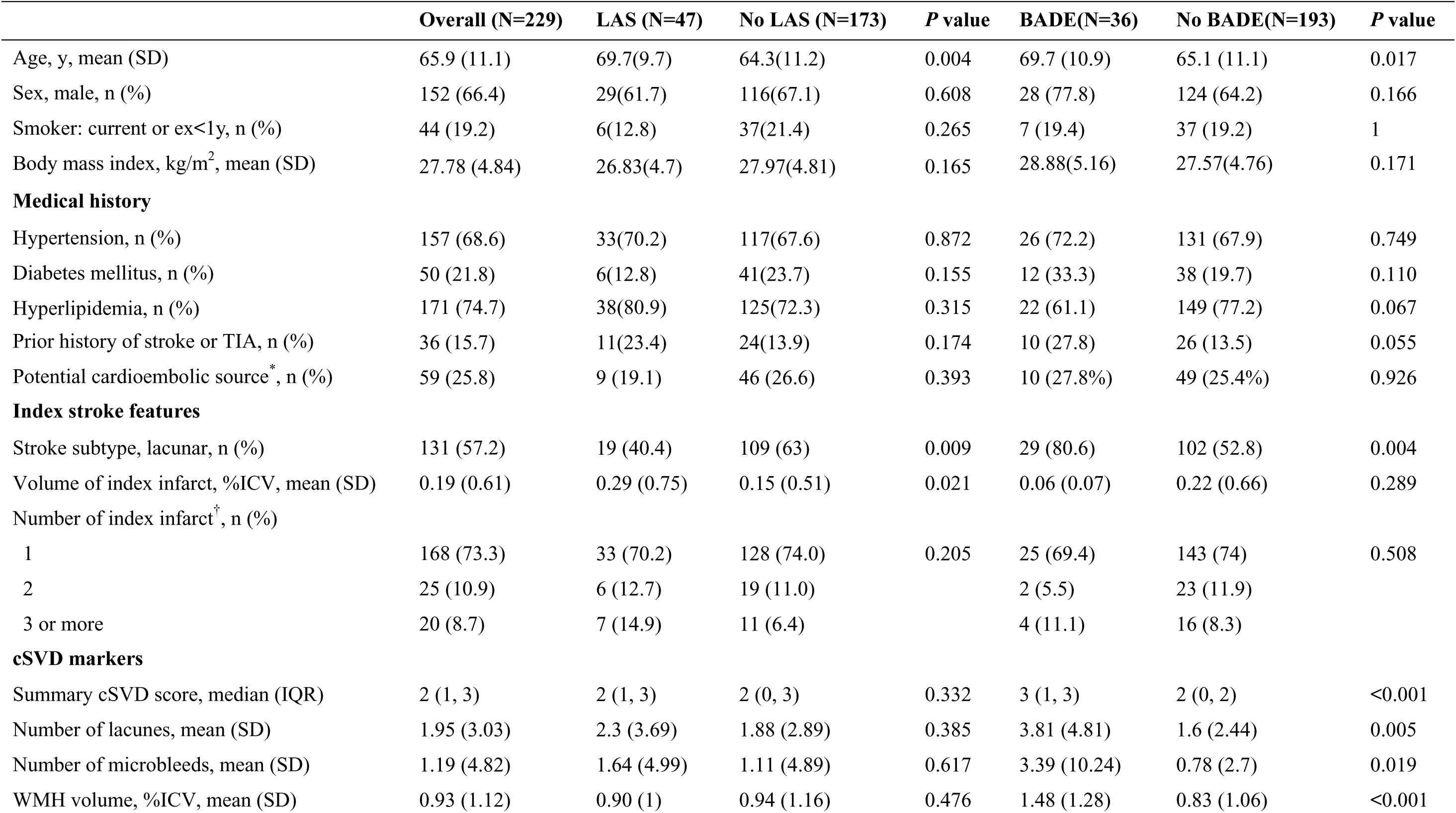
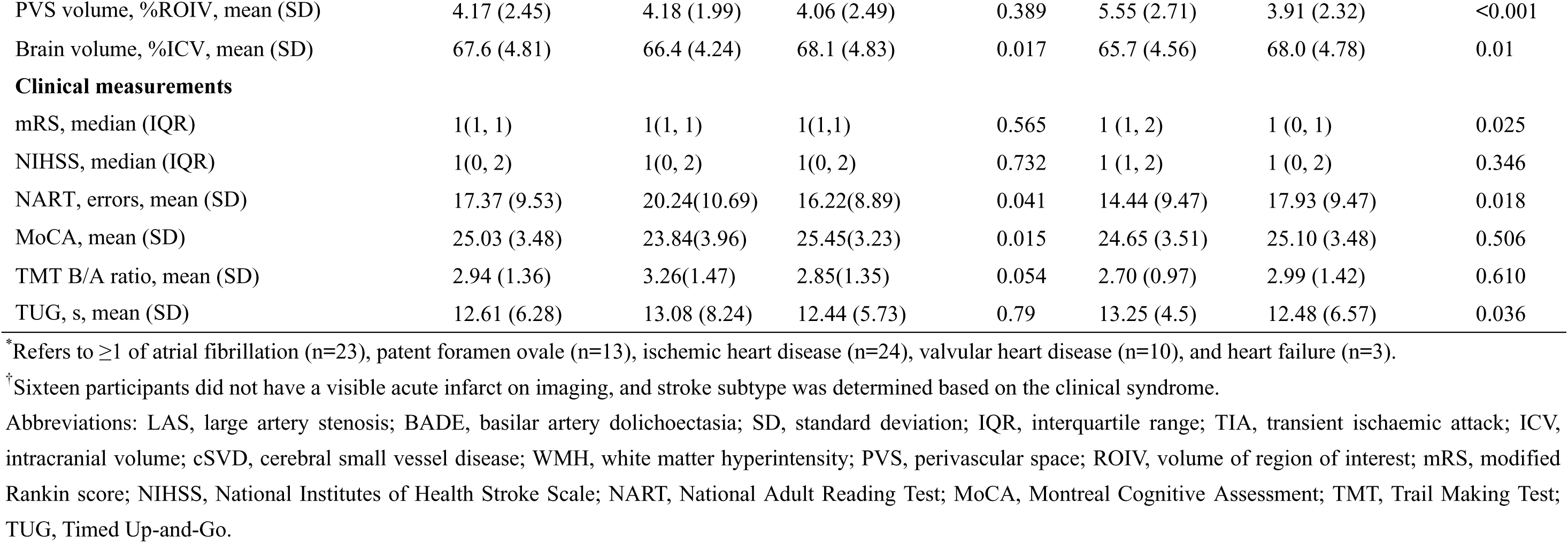
Baseline characteristics of the study population.

## Associations with index stroke

LAS was more prevalent in non-lacunar than lacunar stroke (28/98 [28.5%] vs 19/131 [14.5%]). Among LAS patients, most non-lacunar (22/28, 78.6%) but only 36.8% (7/19) of lacunar strokes involved the territory of the stenotic artery (*P*=0.01; Figure 2A). After adjustment for age, sex, and vascular risk factors, LAS remained less common in lacunar stroke (OR, 0.49; 95% CI, 0.23-0.99) and was associated with a greater number of index infarcts (β, 0.54; 95% CI, 0.14-0.93), whereas its correlation with infarct volume was not retained (Table 2). BADE was more frequent in lacunar than non-lacunar stroke (29/131 [22.1%] vs 7/98 [7.1%]) and remained independently associated with higher odds of lacunar stroke after adjustment (OR, 4.67; 95% CI, 1.87-13.14).

**Figure 2.**
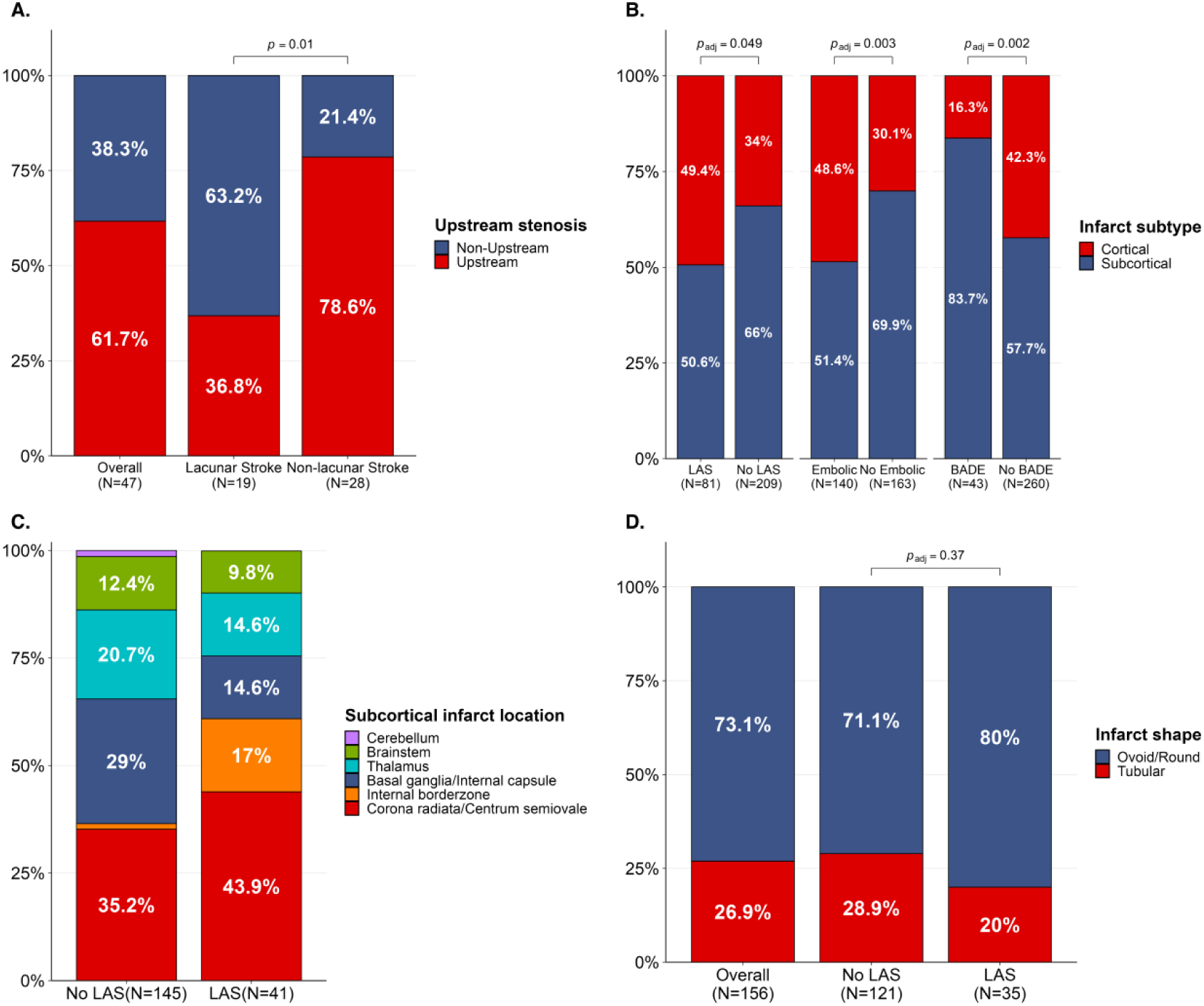
Distributions and anatomical features of index infarcts. (A) Territorial concordance between index strokes and upstream stenosis in the per-person analysis among 47 patients with LAS. (B) Distribution of 303 infarcts at index stroke in the per-infarct analysis, stratified by the presence or absence of LAS, any potential embolic source, and BADE (*P* values from Rao-Scott cluster-adjusted χ² tests). (C) Distribution of anatomical locations of 186 subcortical infarcts at index stroke in the per-infarct analysis, stratified by the presence of LAS. (D) Distribution of shapes of subcortical infarcts in the per-infarct analysis, stratified by the presence of LAS (*P* value from Rao-Scott cluster-adjusted χ² tests). Abbreviations: LAS, large artery stenosis; BADE, basilar artery dolichoectasia.

**Table 2.** Multivariable regression analyses of associations between LAS, BADE, ICAS, and any potential embolic source with index stroke and incident infarcts.

|  | LAS |  | BADE |  | ICAS* |  | Potential embolic source† |  |
| --- | --- | --- | --- | --- | --- | --- | --- | --- |
|  | Effect size (95%CI) ‡ | P value | Effect size (95%CI) ‡ | P value | Effect size (95%CI) ‡ | P value | Effect size (95%CI) ‡ | P value |
| **Index stroke** |  |  |  |  |  |  |  |  |
| Subtype: lacunar vs non-lacunar | OR: 0.49 (0.23, 0.99) | 0.049 | OR: 4.67 (1.87, 13.14) | 0.002 | OR: 0.38 (0.17, 0.83) | 0.016 | OR: 0.47 (0.26, 0.83) | 0.010 |
| Volume of index infarct, %ICV | β: 0.16 (-0.13, 0.45) | 0.286 | β: -0.05 (-0.39, 0.29) | 0.776 | β: 0.24 (-0.08, 0.56) | 0.144 | β: 0.08 (-0.16, 0.33) | 0.498 |
| Number of index infarct | β: 0.54 (0.14, 0.93) | 0.008 | β: -0.13 (-0.56, 0.31) | 0.567 | β: 0.68 (0.29, 1.07) | <0.001 | β: 0.25 (-0.07, 0.56) | 0.129 |
| **Incident infarct** |  |  |  |  |  |  |  |  |
| Presence of incident infarct | OR: 0.88 (0.37, 1.97) | 0.761 | OR: 2.29 (1.01, 5.14) | 0.044 | OR: 1.08 (0.43, 2.53) | 0.867 | OR: 1.30 (0.68, 2.5) | 0.426 |
| Number of incident infarct | β: 0.17 (-0.26, 0.60) | 0.434 | β: 0.97 (0.51, 1.42) | <0.001 | β: 0.26 (-0.18, 0.69) | 0.246 | β: 0.16 (-0.19, 0.5) | 0.369 |
| Symptomatic recurrence | OR: 2.62 (0.91, 7.30) | 0.067 | OR: 0.93 (0.24, 2.99) | 0.912 | OR: 3.50 (1.16, 10.36) | 0.023 | OR: 1.29 (0.50, 3.30) | 0.593 |

\*Analyses involving ICAS were restricted to participants with available intracranial arterial imaging (n = 196)
†Refers to ≥1 of LAS, atrial fibrillation, patent foramen ovale, ischemic heart disease, valvular heart disease, and heart failure.
‡The OR was estimated using binary logistic regression, and the β using linear regression. Models were adjusted for age, sex, and vascular risk factors (smoking, hypertension, diabetes mellitus, hyperlipidemia, and body mass index).
Abbreviations: LAS, large artery stenosis; BADE, basilar artery dolichoectasia; ICAS, intracranial atherosclerotic stenosis; OR, odds ratio; CI, confidence interval; ICV, intracranial volume.

In the per-lesion analysis of 303 index infarcts (186 [61.4%] subcortical, 117 [38.6%] cortical; Table S3), cortical infarcts were more frequent with LAS (49.4% vs 34%, *P*=0.049) and potential embolic source, whereas subcortical were more common with BADE (83.7% vs 57.7%, *P*=0.002; Figure 2B). With LAS, subcortical infarcts more often involved corona radiata, centrum semiovale and internal borderzone, while without LAS they more frequently affected basal ganglia, thalamus, and brainstem (Figure 2C). Infarct shape did not differ by LAS (Figure 2D). Posterior circulation infarcts did not differ by BADE, but brainstem involvement was more common with BADE (16.3% vs 5.8%; Table S4).

## Associations with incident infarcts

Over one year, 130 incident infarcts (97 [74.6%] subcortical, 33 [25.4%] cortical) occurred in 59 participants. At the per-person level, BADE was associated with higher risk (OR, 2.29; 95% CI, 1.01-5.14) and number (β, 0.97; 95% CI, 0.51-1.42) of incident infarcts after adjustment, whereas LAS showed no significant association (Table 2). Recurrent symptomatic stroke or TIA occurred in 22 participants, with a trend toward higher risk with LAS (OR, 2.62; 95% CI, 0.91-7.30; *P*=0.067) and a significant association with ICAS (OR, 3.50; 95% CI, 1.16-10.36; *P*=0.023).

At the per-lesion level, cortical incident infarcts were more frequent with an embolic source (36.8% vs 16.4%), while subcortical were more common with BADE (86% vs 67.5%). Among 26 incident infarcts in 10 LAS patients, all cortical infarcts (5/5) occurred downstream of the stenosis, while most subcortical (15/21, *P*=0.049) did not. Subcortical infarcts predominantly involved cerebral white matter (Figure S3, Table S5).

## Associations with cSVD markers and progression

After adjustment for demographics and vascular risk factors, LAS was not associated with baseline cSVD markers (Table 3). In contrast, BADE was related to multiple markers, including more lacunes (β, 2.31; 95% CI, 1.27-3.36) and microbleeds (β, 3.41; 95% CI, 1.68-5.13), greater WMH (β, 0.26; 95% CI, 0.11-0.41) and PVS (β, 0.10; 95% CI, 0.02-0.18) volumes, and higher summary cSVD score (OR, 2.57; 95% CI, 1.28-5.25). No associations were observed between BADE and brain atrophy. At one year, BADE remained associated with all cSVD markers and additionally with WMH progression (β, 0.15; 95% CI, 0.01-0.29), while LAS was related only to more lacunes.

**Table 3.** Multivariable regression analyses of associations between LAS, BADE, and cSVD markers at baseline and one year.

|  | LAS |  | BADE |  |
| --- | --- | --- | --- | --- |
|  | Effect size (95%CI) | P value | Effect size (95%CI) | P value |
| **Baseline cSVD markers\*** |  |  |  |  |
| Summary cSVD score | OR: 1.08 (0.59, 1.96) | 0.809 | OR: 2.57 (1.28, 5.25) | 0.009 |
| Number of lacunes | $\beta$: 0.75 (-0.25, 1.74) | 0.139 | $\beta$: 2.31 (1.27, 3.36) | <0.001 |
| Number of microbleeds | $\beta$: 0.88 (-0.76, 2.52) | 0.289 | $\beta$: 3.41 (1.68, 5.13) | <0.001 |
| WMH volume, %ICV | $\beta$: -0.03 (-0.17, 0.11) | 0.696 | $\beta$: 0.26 (0.11, 0.41) | <0.001 |
| BG PVS volume, %ROIV | $\beta$: -0.05 (-0.11, 0.01) | 0.125 | $\beta$: 0.09 (0.03, 0.16) | 0.005 |
| CSO PVS volume, %ROIV | $\beta$: -0.02 (-0.11, 0.06) | 0.586 | $\beta$: 0.10 (0.01, 0.20) | 0.034 |
| Total PVS volume, %ROIV | $\beta$: -0.03 (-0.10, 0.05) | 0.494 | $\beta$: 0.10 (0.02, 0.18) | 0.018 |
| Brain volume, %ICV | $\beta$: -0.09 (-1.17, 0.99) | 0.867 | $\beta$: 0.14 (-1.11, 1.40) | 0.822 |
| Deep atrophy score | OR: 1.28 (0.70, 2.35) | 0.422 | OR: 1.74 (0.86, 3.54) | 0.124 |
| Superficial atrophy score | OR: 0.91 (0.49, 1.70) | 0.755 | OR: 1.18 (0.56, 2.46) | 0.666 |
| **One-year cSVD markers†** |  |  |  |  |
| Number of lacunes | $\beta$: 0.84 (0.04, 1.64) | 0.040 | $\beta$: 1.56 (0.72, 2.41) | <0.001 |
| Number of microbleeds | $\beta$: 0.92 (-0.97, 2.81) | 0.338 | $\beta$: 2.98 (1.01, 4.96) | 0.003 |
| WMH volume, %ICV | $\beta$: -0.08 (-0.20, 0.03) | 0.161 | $\beta$: 0.16 (0.04, 0.29) | 0.011 |
| WMH progression, %ICV | $\beta$: -0.04 (-0.17, 0.09) | 0.560 | $\beta$: 0.15 (0.01, 0.29) | 0.032 |
| BG PVS volume, %ROIV | $\beta$: -0.05 (-0.10, 0.01) | 0.082 | $\beta$: 0.08 (0.02, 0.14) | 0.006 |
| CSO PVS volume, %ROIV | $\beta$: -0.04 (-0.13, 0.04) | 0.320 | $\beta$: 0.11 (0.02, 0.21) | 0.023 |
| Total PVS volume, %ROIV | $\beta$: -0.04 (-0.12, 0.03) | 0.228 | $\beta$: 0.10 (0.02, 0.18) | 0.011 |
| Brain volume, %ICV | $\beta$: -0.29 (-1.42, 0.83) | 0.611 | $\beta$: 0.09 (-1.18, 1.36) | 0.894 |
| Deep atrophy score | OR: 1.38 (0.71, 2.67) | 0.336 | OR: 2.16 (1.03, 4.56) | 0.043 |
| Superficial atrophy score | OR: 0.95(0.49, 1.84) | 0.880 | OR: 1.36 (0.63, 2.97) | 0.435 |

\*Models for baseline cSVD markers were adjusted for age, sex, and vascular risk factors. For brain volume, deep atrophy score, and superficial atrophy score, models were additionally adjusted for the number of lacunes at baseline.
†Models for one-year cSVD markers were adjusted for the same covariates as above, additionally for baseline cSVD score. For brain volume, deep atrophy score, and superficial atrophy score, models were additionally adjusted for the number of lacunes at one year.
$\beta$ coefficients were estimated using linear regression, and ORs using ordinal logistic regression.
Abbreviations: LAS, large artery stenosis; BADE, basilar artery dolichoectasia; cSVD, cerebral small vessel disease; OR, odds ratio; CI, confidence interval; WMH, white matter hyperintensity; ICV, intracranial volume; BG PVS, basal ganglia perivascular space; CSO PVS, centrum semiovale perivascular space; ROIV, volume of region of interest.

## Associations with clinical outcomes

In univariate analyses, LAS was linked to lower MoCA scores, and BADE to higher mRS and longer TUG times (Table 1). However, in multivariable models, neither LAS nor BADE remained significantly associated with functional (mRS), cognitive (MoCA, TMT B/A ratio), or mobility (TUG) outcomes at baseline or one year (Table S6).

## Sensitivity analysis

Associations of ICAS and potential embolic source with index and incident infarcts were consistent with those of LAS (Table 2). Additional adjustment for baseline cSVD score did not alter these associations (Table S7). In models examining cSVD markers, results remained broadly consistent when BADE was modeled using continuous arterial diameters and when LAS was replaced by ICAS, potential embolic source, or atrial fibrillation (Table S8). In addition, inclusion of a multiplicative interaction term between LAS and BADE in both infarct and cSVD models did not indicate evidence of effect modification (Table S9).

## Systematic review

Findings from the systematic review are detailed in the Supplemental Material (Figure S4). Across 27 studies comprising 9515 patients with lacunar stroke, the pooled prevalence of ipsilateral LAS was 11% (95% CI, 8-15%; Table S10 and Figure S5A-C), higher in Asian than Caucasian cohorts (19% vs. 9%, Figure S5D-E). In 13 studies directly comparing ipsilateral and contralateral LAS, no significant difference was found (pooled risk difference, 0.01; 95% CI, –0.04 to 0.06; Figure S5F), providing little evidence for a causal relationship. In community cohorts, LAS often correlated with lacunes and WMH, but findings in stroke cohorts were inconsistent (Table S11-S14). Even when associations existed, no consistent laterality was observed between LAS and lacunes or WMH (Table S12, S14). Most studies reported no association between LAS and microbleeds (Table S15) or PVS (Table S16). Fifteen cross-sectional studies assessing dolichoectasia showed associations with lacunar stroke and multiple cSVD markers, which were further supported by a meta-analysis (Table S17, Figure S6).

## Discussion

In this prospective cohort of patients with lacunar stroke enriched for cSVD features and non-lacunar controls, cranial artery stenosis and dolichoectasia showed distinct associations with stroke subtype, cSVD and its progression. LAS was associated with non-lacunar stroke and a higher risk of symptomatic recurrent stroke or TIA but was uncommon in lacunar stroke and unrelated to MRI-defined incident infarcts or cSVD markers. Similar patterns were observed for ICAS and embolic source. In contrast, BADE was more frequent in lacunar subtype and, together with larger arterial diameters, strongly associated with all cSVD markers except atrophy. BADE was also associated with more incident infarcts on follow-up MRI most of which were subcortical, though not with symptomatic recurrence. Neither LAS nor BADE independently predicted functional, cognitive, or mobility outcomes at baseline or one year.

The prevalence of LAS (20.5%) in our cohort was comparable to previous reports in minor stroke.^27,28^ LAS and other embolic sources were more commonly associated with non-lacunar than lacunar stroke, across both index and incident infarcts and in per-person and per-lesion analyses. While non-lacunar strokes often aligned with proximal stenosis, lacunar strokes showed weaker territorial concordance, suggesting non-causal coexistence. This is supported by the Secondary Prevention of Small Subcortical Strokes trial, where 17.3% of patients with lacunar stroke had ICAS but only 1.8% had relevant ipsilateral stenosis.^29,30^ Similarly, our meta-analysis demonstrated no difference in stenosis prevalence between ipsilateral and contralateral arteries in lacunar stroke. These findings are consistent with Fisher’s seminal pathological observations. Although all four patients that he examined had severe atherosclerosis at the patient level, reflecting shared vascular risk factors, lesion-level analysis showed that the majority of lacunar lesions were attributable to intrinsic small vessel disease, termed ‘segmental arterial disorganization’ (40/50 [80%]), whereas parent artery atheroma directly occluding the arteriole supplying the lacunar lesion was rare (3/50 [6%]).^7^

In LAS patients, lacunar-appearing infarcts were more often located in internal borderzone rather than deep perforator territories and were frequently multiple. This pattern supports that proximal stenosis may contribute through embolic or hemodynamic mechanisms, particularly when infarcts are multifocal and unilateral, rather than by direct perforator occlusion.^11^ These findings highlight the need for individualized etiologic assessment. In typical lacunar stroke, where intrinsic cSVD predominates, non-selective embolic screening may have limited diagnostic value, although it remains essential to exclude treatable large artery or cardioembolic causes when suspected.

Although LAS/ICAS was linked to a higher risk of symptomatic recurrent stroke or TIA, consistent with previous studies,^27,31^ it was unrelated to new MRI-detected infarcts, which were mostly covert, small, and subcortical. More than one-quarter of participants developed these incident infarcts despite guideline-based secondary stroke prevention, and they were strongly associated with baseline cSVD burden,^18^ supporting cSVD as a progressive, largely non-atheromatous process. Despite the presence of shared vascular risk factors across stroke subtypes, our findings suggest distinct downstream pathophysiological mechanisms, with atherosclerosis predominating in non-lacunar stroke and intrinsic microvascular disease underlying lacunar (cSVD-related) stroke. Current secondary prevention strategies for lacunar stroke and cSVD largely mirror those used for large-artery atherosclerotic disease, including vascular risk factor control, antihypertensive, antiplatelet, and lipid-lowering treatment. While these approaches remain important and should not be discounted, accumulating evidence, consistent with our results, suggests that guideline-based secondary stroke prevention including antiplatelet therapy and statins has limited efficacy in preventing progression of cSVD-related brain damage.^32,33^ Collectively, these findings emphasize that in lacunar stroke with substantial cSVD burden, management should target microvascular pathology, for example through therapies aimed at improving microvascular function, rather than relying solely on atherosclerosis-focused approaches.^34–37^

Although some community-based studies have reported associations between LAS and certain cSVD markers, findings in stroke populations were inconsistent. In our mild stroke cohort, LAS showed no such association. This discrepancy may reflect methodological differences: inclusion of a well-defined non-lacunar control group and rigorous adjustment for vascular risk factors and baseline cSVD enabled distinction between co-association and causation. Many prior studies lacked appropriate controls, inadequately adjusted for confounders, and included heterogeneous populations or disease stages, likely explaining the inconsistent results.

Consistent with previous studies,^4^ BADE was strongly associated with multiple cSVD features, including a four-fold increased odds of lacunar stroke, greater cSVD burden and progression, and more incident subcortical infarcts. Several mechanisms may underlie this association. First, shared genetic susceptibility may contribute: mutations in basement membrane protein genes (e.g., *COL4A1* and *COL4A2*) have been identified in patients with dolichoectasia and cSVD, and showed associations with WMH in genome wide association studies.^38,39^ These findings support a genetic continuum linking large artery widening and intrinsic small vessel arteriopathy. Second, both conditions may reflect connective tissue abnormalities of the vessel wall. Fisher and others described microarteriolar dilatation, elastic lamina fragmentation, and medial rarefaction in cSVD, possibly due to extracellular matrix imbalance and dysregulated metalloproteinase activity, leading to arterial weakening and widening in large vessels and segmental disorganization in small arterioles.^7,40–42^ Third, widened or tortuous arteries may exert mechanical stress on perforating arterioles, disrupting local flow and promoting microvascular injury.^43^ Supporting this, we observed more pontine infarcts in BADE.

To our knowledge, this is the first study to evaluate two distinct large artery pathologies within a single, well-characterized cohort, controlling for stroke subtype and medication, enabling direct comparison of their associations with stroke subtypes and cSVD. Key strengths include its prospective design, standardized and volumetric assessment of comprehensive cSVD features at both patient and lesion levels, and blinded evaluation of large artery characteristics to all clinical, imaging, and outcome data. The findings were further contextualized within an updated systematic review and meta-analysis. Several limitations should be acknowledged. First, ICAS was evaluated using arterial-phase DCE-MRI rather than dedicated angiography. Although potentially less sensitive to mild stenosis, the slow-injection technique provides high-quality luminal visualization comparable to conventional bolus MR angiography, making it unlikely to miss moderate or severe stenosis. Second, the lack of vessel wall imaging limited direct visualization of intramural plaque at perforator origins. Third, the single-center sample limits generalizability, underscoring the need for larger multicenter studies to validate and extend these findings.

## Conclusions

Large artery pathologies exhibit divergent etiopathological implications for stroke subtype and cSVD. Dolichoectasia reflects a cSVD-related phenotype, while LAS was uncommon in lacunar stroke and unrelated to cSVD markers. These findings support that lacunar stroke and other cSVD manifestations are predominantly driven by intrinsic, non-atheromatous microvascular pathology, highlighting the need for mechanism-based diagnostic and therapeutic strategies beyond conventional approaches targeting atheroma or cardioembolism.

### Sources of funding

This study was funded by the UK Dementia Research Institute, which receives its funding from DRI Ltd. funded by the UK Medical Research Council, Alzheimer’s Society and Alzheimer’s Research UK; the Fondation Leducq Network for the Study of Perivascular Spaces in Small Vessel Disease (16 CVD 05); Stroke Association ‘Small Vessel Disease-Spotlight on Symptoms (SVD-SOS)’ (SAPG 19\1000680); European Union Horizon 2020 (No. 666881, ‘SVDs@Target’); British Heart Foundation Edinburgh Centre for Research Excellence (RE/18/5/34216); The Mrs Gladys Row Fogo Charitable Trust Centre for Research into Aging and the Brain. F.H. received funding from National Natural Science Foundation of China (No.82271368, No.82571510), and the National High-Level Hospital Clinical Research Funding (2025-PUMCH-A-151); U.C. was funded by the Chief Scientist Office of Scotland Clinical Academic Fellowship (CAF/18/08) and Stroke Association Princess Margaret Research Development Fellowship (2018); C.A.R. received funding from Mexican Council of Humanities Science and Technology (CONAHCYT, 2021-000007-01EXTF-00234), Anne Rowling Regenerative Neurology Clinic, and Row Fogo Charitable Trust; M.J.T received funding from NHS Lothian Research and Development Office; D.J.G was funded by the Wellcome Trust Translational Neuroscience Ph.D. programme (224912/ Z/21/Z); M.S.S was funded by a Stroke Association Post-Doctoral Fellowships (SAPDF 23/100007); Y.C. was supported by the Postdoctor Research Fund of West China Hospital, Sichuan University (2024HXBH041) and China Scholarship Council (CSC202006240281); A.C.C.J. received funding from the Alzheimer’s Society (AS-CP-18b-001); F.N.D. was funded by the Stroke Association-Garfield Weston Foundation Senior Clinical Lectureship (TSALECT 2015/04), NHS Research Scotland and the Agnes Parry Bequest; The 3T MRI scanner is supported by the Scottish Funding Council through the Scottish Imaging Network, A Platform for Scientific Excellence (SINAPSE) Collaboration; the Wellcome Trust (104916/Z/14/Z), Dunhill Trust (R380R/1114), Edinburgh and Lothians Health Foundation (2012/17), Muir Maxwell Research Fund and the University of Edinburgh.

### Disclosures

None reported.

### Supplemental Material

Supplemental Methods

Figure S1-S6

Table S1-S17

## Non-standard Abbreviations and Acronyms

BA: basilar artery
BADE: basilar artery dolichoectasia
cSVD: cerebral small vessel disease
DCE: dynamic contrast-enhanced
DWI: diffusion-weighted imaging
FLAIR: fluid attenuated inversion recovery
ICA: internal carotid arteries
ICAS: intracranial atherosclerotic stenosis
ICV: intracranial volume
LAS: Large artery stenosis
MCA: middle cerebral arteries
MoCA: Montreal Cognitive Assessment
mRS: modified Rankin scale
NART: National Adult Reading Test
OR: odds ratio
PVS: perivascular space
ROIV: volume of region of interest
TIA: transient ischemic attack
TMT: Trail Making Test
TUG: Timed Up-and-Go
WMH: white matter hyperintensity

## Notes

### Competing Interest Statement

The authors have declared no competing interest.

## References

1. Gutierrez J, Turan TN, Hoh BL, Chimowitz MI. Intracranial atherosclerotic stenosis: risk factors, diagnosis, and treatment. Lancet Neurol. 2022;21:355–368. doi: 10.1016/S1474-4422(21)00376-8

2. Pico F, Labreuche J, Amarenco P. Pathophysiology, presentation, prognosis, and management of intracranial arterial dolichoectasia. Lancet Neurol. 2015;14:833–845. doi: 10.1016/s1474-4422(15)00089-7

3. Thiankhaw K, Ozkan H, Ambler G, Werring DJ. Relationships between intracranial arterial dolichoectasia and small vessel disease in patients with ischaemic stroke: a systematic review and meta-analysis. J Neurol. 2024;271:772–781. doi: 10.1007/s00415-023-12094-2

4. Zhang DP, Yin S, Zhang HL, Li D, Song B, Liang JX. Association between Intracranial Arterial Dolichoectasia and Cerebral Small Vessel Disease and Its Underlying Mechanisms. J Stroke. 2020;22:173–184. doi: 10.5853/jos.2019.02985

5. Brisset M, Boutouyrie P, Pico F, Zhu Y, Zureik M, Schilling S, Dufouil C, Mazoyer B, Laurent S, Tzourio C, et al. Large-vessel correlates of cerebral small-vessel disease. Neurology. 2013;80:662–669. doi: 10.1212/WNL.0b013e318281ccc2

6. Regenhardt RW, Das AS, Lo EH, Caplan LR. Advances in Understanding the Pathophysiology of Lacunar Stroke: A Review. JAMA Neurol. 2018;75:1273–1281. doi: 10.1001/jamaneurol.2018.1073

7. Fisher CM. The arterial lesions underlying lacunes. Acta Neuropathol. 1968;12:1–15. doi: 10.1007/BF00685305

8. Adachi T, Kobayashi S, Yamaguchi S, Okada K. MRI findings of small subcortical “lacunar-like” infarction resulting from large vessel disease. J Neurol. 2000;247:280–285. doi: 10.1007/s004150050584

9. Jackson CA, Hutchison A, Dennis MS, Wardlaw JM, Lindgren A, Norrving B, Anderson CS, Hankey GJ, Jamrozik K, Appelros P, et al. Differing risk factor profiles of ischemic stroke subtypes: evidence for a distinct lacunar arteriopathy? Stroke. 2010;41:624–629. doi: 10.1161/strokeaha.109.558809

10. Mead GE, Lewis SC, Wardlaw JM, Dennis MS, Warlow CP. Severe ipsilateral carotid stenosis and middle cerebral artery disease in lacunar ischaemic stroke: innocent bystanders? J Neurol. 2002;249:266–271. doi: 10.1007/s004150200003

11. Tejada J, Díez-Tejedor E, Hernández-Echebarría L, Balboa O. Does a relationship exist between carotid stenosis and lacunar infarction? Stroke. 2003;34:1404–1409. doi: 10.1161/01.Str.0000072520.53106.8c

12. Clancy U, Garcia DJ, Stringer MS, Thrippleton MJ, Valdes-Hernandez MC, Wiseman S, Hamilton OK, Chappell FM, Brown R, Blair GW, et al. Rationale and design of a longitudinal study of cerebral small vessel diseases, clinical and imaging outcomes in patients presenting with mild ischaemic stroke: Mild Stroke Study 3. Eur Stroke J. 2021;6:81–88. doi: 10.1177/2396987320929617

13. Bamford J, Sandercock P, Dennis M, Burn J, Warlow C. Classification and natural history of clinically identifiable subtypes of cerebral infarction. Lancet. 1991;337:1521–1526. doi: 10.1016/0140-6736(91)93206-o

14. Potter G, Doubal F, Jackson C, Sudlow C, Dennis M, Wardlaw J. Associations of clinical stroke misclassification (’clinical-imaging dissociation’) in acute ischemic stroke. Cerebrovasc Dis. 2010;29:395–402. doi: 10.1159/000286342

15. Makin SD, Doubal FN, Shuler K, Chappell FM, Staals J, Dennis MS, Wardlaw JM. The impact of early-life intelligence quotient on post stroke cognitive impairment. Eur Stroke J. 2018;3:145–156. doi: 10.1177/2396987317750517

16. Nasreddine ZS, Phillips NA, Bedirian V, Charbonneau S, Whitehead V, Collin I, Cummings JL, Chertkow H. The Montreal Cognitive Assessment, MoCA: a brief screening tool for mild cognitive impairment. J Am Geriatr Soc. 2005;53:695–699. doi: 10.1111/j.1532-5415.2005.53221.x

17. Bowie CR, Harvey PD. Administration and interpretation of the Trail Making Test. Nat Protoc. 2006;1:2277–2281. doi: 10.1038/nprot.2006.390

18. Clancy U, Arteaga-Reyes C, Jaime Garcia D, Hewins W, Locherty R, Valdes Hernandez MDC, Wiseman SJ, Stringer MS, Thrippleton M, Chappell FM, et al. Incident Infarcts in Patients With Stroke and Cerebral Small Vessel Disease: Frequency and Relation to Clinical Outcomes. Neurology. 2024;103:e209750. doi: 10.1212/WNL.0000000000209750

19. Duering M, Biessels GJ, Brodtmann A, Chen C, Cordonnier C, de Leeuw FE, Debette S, Frayne R, Jouvent E, Rost NS, et al. Neuroimaging standards for research into small vessel disease—advances since 2013. Lancet Neurol. 2023;22:602–618. doi: 10.1016/s1474-4422(23)00131-x

20. Staals J MS, Doubal FN, Dennis MS, Wardlaw JM. Stroke subtype, vascular risk factors, and total MRI brain small-vessel disease burden. Neurology. 2014;83:1228–1234.

21. Farrell C, Chappell F, Armitage PA, Keston P, Maclullich A, Shenkin S, Wardlaw JM. Development and initial testing of normal reference MR images for the brain at ages 65-70 and 75-80 years. Eur Radiol. 2009;19:177–183. doi: 10.1007/s00330-008-1119-2

22. Valdés Hernández MdC, Ballerini L, Glatz A, Aribisala BS, Bastin ME, Dickie DA, Duarte Coello R, Munoz Maniega S, Wardlaw JM [database online].

23. Valdes Hernandez Mdel C, Armitage PA, Thrippleton MJ, Chappell F, Sandeman E, Munoz Maniega S, Shuler K, Wardlaw JM. Rationale, design and methodology of the image analysis protocol for studies of patients with cerebral small vessel disease and mild stroke. Brain Behav. 2015;5:e00415. doi: 10.1002/brb3.415

24. Samuels OB, Joseph GJ, Lynn MJ, Smith HA, Chimowitz MI. A standardized method for measuring intracranial arterial stenosis. AJNR Am J Neuroradiol. 2000;21:643–646.

25. Barnett HJ, Taylor DW, Eliasziw M, Fox AJ, Ferguson GG, Haynes RB, Rankin RN, Clagett GP, Hachinski VC, Sackett DL, et al. Benefit of carotid endarterectomy in patients with symptomatic moderate or severe stenosis. North American Symptomatic Carotid Endarterectomy Trial Collaborators. N Engl J Med. 1998;339:1415–1425. doi: 10.1056/NEJM199811123392002

26. Smoker WR, Price MJ, Keyes WD, Corbett JJ, Gentry LR. High-resolution computed tomography of the basilar artery: 1. Normal size and position. AJNR Am J Neuroradiol. 1986;7:55–60.

27. Amarenco P, Lavallee PC, Labreuche J, Albers GW, Bornstein NM, Canhao P, Caplan LR, Donnan GA, Ferro JM, Hennerici MG, et al. One-Year Risk of Stroke after Transient Ischemic Attack or Minor Stroke. N Engl J Med. 2016;374:1533–1542. doi: 10.1056/NEJMoa1412981

28. Hurford R, Wolters FJ, Li L, Lau KK, Kuker W, Rothwell PM, Oxford Vascular Study Phenotyped C. Prevalence, predictors, and prognosis of symptomatic intracranial stenosis in patients with transient ischaemic attack or minor stroke: a population-based cohort study. Lancet Neurol. 2020;19:413–421. doi: 10.1016/S1474-4422(20)30079-X

29. Asdaghi N, Pearce LA, Nakajima M, Field TS, Bazan C, Cermeno F, McClure LA, Anderson DC, Hart RG, Benavente OR. Clinical correlates of infarct shape and volume in lacunar strokes: the Secondary Prevention of Small Subcortical Strokes trial. Stroke. 2014;45:2952–2958. doi: 10.1161/strokeaha.114.005211

30. Palacio S, McClure LA, Benavente OR, Bazan C, 3rd, Pergola P, Hart RG. Lacunar strokes in patients with diabetes mellitus: risk factors, infarct location, and prognosis: the secondary prevention of small subcortical strokes study. Stroke. 2014;45:2689–2694. doi: 10.1161/strokeaha.114.005018

31. Lovett JK, Coull AJ, Rothwell PM. Early risk of recurrence by subtype of ischemic stroke in population-based incidence studies. Neurology. 2004;62:569–573. doi: 10.1212/01.wnl.0000110311.09970.83

32. Wardlaw JM, Chabriat H, de Leeuw FE, Debette S, Dichgans M, Doubal F, Jokinen H, Katsanos AH, Ornello R, Pantoni L, et al. European stroke organisation (ESO) guideline on cerebral small vessel disease, part 2, lacunar ischaemic stroke. Eur Stroke J. 2024;9:5–68. doi: 10.1177/23969873231219416

33. Wardlaw JM, Debette S, Jokinen H, De Leeuw FE, Pantoni L, Chabriat H, Staals J, Doubal F, Rudilosso S, Eppinger S, et al. ESO Guideline on covert cerebral small vessel disease. Eur Stroke J. 2021;6:CXI–CLXII. doi: 10.1177/23969873211012132

34. Bath PM, Wardlaw JM. Pharmacological treatment and prevention of cerebral small vessel disease: a review of potential interventions. Int J Stroke. 2015;10:469–478. doi: 10.1111/ijs.12466

35. Brown RB, Tozer DJ, Loubiere L, Harshfield EL, Hong YT, Fryer TD, Williams GB, Graves MJ, Aigbirhio FI, O’Brien JT, et al. MINocyclinE to Reduce inflammation and blood-brain barrier leakage in small Vessel diseAse (MINERVA): A phase II, randomized, double-blind, placebo-controlled experimental medicine trial. Alzheimers Dement. 2024;20:3852–3863. doi: 10.1002/alz.13830

36. Kopczak A, Stringer MS, van den Brink H, Kerkhofs D, Blair GW, van Dinther M, Reyes CA, Garcia DJ, Onkenhout L, Wartolowska KA, et al. Effect of blood pressure-lowering agents on microvascular function in people with small vessel diseases (TREAT-SVDs): a multicentre, open-label, randomised, crossover trial. Lancet Neurol. 2023;22:991–1004. doi: 10.1016/S1474-4422(23)00293-4

37. Wardlaw JM, Woodhouse LJ, Mhlanga, II, Oatey K, Heye AK, Bamford J, Cvoro V, Doubal FN, England T, Hassan A, et al. Isosorbide Mononitrate and Cilostazol Treatment in Patients With Symptomatic Cerebral Small Vessel Disease: The Lacunar Intervention Trial-2 (LACI-2) Randomized Clinical Trial. JAMA Neurol. 2023;80:682–692. doi: 10.1001/jamaneurol.2023.1526

38. Rannikmae K, Sivakumaran V, Millar H, Malik R, Anderson CD, Chong M, Dave T, Falcone GJ, Fernandez-Cadenas I, Jimenez-Conde J, et al. COL4A2 is associated with lacunar ischemic stroke and deep ICH: Meta-analyses among 21,500 cases and 40,600 controls. Neurology. 2017;89:1829–1839. doi: 10.1212/WNL.0000000000004560

39. Renard D, Mine M, Pipiras E, Labauge P, Delahaye A, Benzacken B, Tournier-Lasserve E. Cerebral small-vessel disease associated with COL4A1 and COL4A2 gene duplications. Neurology. 2014;83:1029–1031. doi: 10.1212/WNL.0000000000000769

40. Pico F, Jacob MP, Labreuche J, Soufir N, Touboul PJ, Benessiano J, Cambien F, Grandchamp B, Michel JB, Amarenco P. Matrix metalloproteinase-3 and intracranial arterial dolichoectasia. Ann Neurol. 2010;67:508–515. doi: 10.1002/ana.21922

41. Pico F, Labreuche J, Seilhean D, Duyckaerts C, Hauw JJ, Amarenco P. Association of small-vessel disease with dilatative arteriopathy of the brain: neuropathologic evidence. Stroke. 2007;38:1197–1202. doi: 10.1161/01.STR.0000259708.05806.76

42. Sho E, Sho M, Singh TM, Nanjo H, Komatsu M, Xu C, Masuda H, Zarins CK. Arterial enlargement in response to high flow requires early expression of matrix metalloproteinases to degrade extracellular matrix. Exp Mol Pathol. 2002;73:142–153. doi: 10.1006/exmp.2002.2457

43. Kumral E, Kisabay A, Atac C, Kaya C, Calli C. The mechanism of ischemic stroke in patients with dolichoectatic basilar artery. Eur J Neurol. 2005;12:437–444. doi: 10.1111/j.1468-1331.2005.00993.x

